# Potential impact of seasonal forcing on a SARS-CoV-2 pandemic

**DOI:** 10.1101/2020.02.13.20022806

**Authors:** Richard A. Neher, Robert Dyrdak, Valentin Druelle, Emma B. Hodcroft, Jan Albert

**Affiliations:** Biozentrum, University of Basel, Basel, Switzerland; Swiss Institute of Bioinformatics, Basel, Switzerland; Department of Clinical Microbiology, Karolinska University Hospital, Stockholm, Sweden; Department of Microbiology, Tumor and Cell Biology, Karolinska Institute, Stockholm, Sweden

**Author notes:** Correspondence to: Richard Neher, Biozentrum, Klingelbergstrasse 70, 4056, Basel, Switzerland.

## Abstract

A novel coronavirus (SARS-CoV-2) first detected in Wuhan, China, has spread rapidly since December 2019, causing more than 80,000 confirmed infections and 2,700 fatalities (as of Feb 27, 2020). Imported cases and transmission clusters of various sizes have been reported globally suggesting a pandemic is likely.

Here, we explore how seasonal variation in transmissibility could modulate a SARS-CoV-2 pandemic. Data from routine diagnostics show a strong and consistent seasonal variation of the four endemic coronaviruses (229E, HKU1, NL63, OC43) and we parameterize our model for SARS-CoV-2 using these data. The model allows for many subpopulations of different size with variable parameters. Simulations of different scenarios show that plausible parameters result in a small peak in early 2020 in temperate regions of the Northern Hemisphere and a larger peak in winter 2020/2021. Variation in transmission and migration rates can result in substantial variation in prevalence between regions.

While the uncertainty in parameters is large, the scenarios we explore show that transient reductions in the incidence rate might be due to a combination of seasonal variation and infection control efforts but do not necessarily mean the epidemic is contained. Seasonal forcing on SARS-CoV-2 should thus be taken into account in the further monitoring of the global transmission. The likely aggregated effect of seasonal variation, infection control measures, and transmission rate variation is a prolonged pandemic wave with lower prevalence at any given time, thereby providing a window of opportunity for better preparation of health care systems.

---

On Jan 30, 2020, the World Health Organisation (WHO) declared the spread of a new coronavirus, SARS-CoV-2 (Gorbalenya, 2020), as a public health emergency of international concern (WHO Emergency Committee, 2020c). The virus was first identified in patients with pneumonia in the city of Wuhan in the Hubei province, China, in December 2019 (Liangjun *et al*., 2020). The clinical presentation of the illness caused by SARS-CoV-2, called COVID-19, appears to range from mild or asymptomatic to severe and fatal respiratory illness (WHO Emergency Committee, 2020a), but the exact spectrum of disease presentation is still unclear. The potential for global spread, i.e. a pandemic, of SARS-CoV-2 is currently not known, but the virus has spread at an alarming rate in Wuhan, the epicenter of the outbreak. Furthermore, the virus has spread to all provinces of China and small clusters of local spread have been reported from several countries, e.g. Singapore, Germany, and the UK (Rothe *et al*., 2020; Singapore Ministry of Health, 2020; WHO Emergency Committee, 2020a).

The basic reproduction number (*R*_0_), which describes the average number of new infections per infected SARS-CoV-2 case, has been estimated to be around *R*_0_ = 2 *-* 3 (2.2 with 90% high density interval 1.4–3.8 (Riou and Althaus, 2020) or 2.7 with a 95% CrI of 2.47–2.86 (Wu *et al*., 2020)). Higher estimates have also been reported (Sanche *et al*., 2020; Yang *et al*., 2020). Importantly, *R*_0_ is not a biological constant for a pathogen, but is affected by factors such as environmental conditions and the behaviour of infected individuals. One such environmental factor is climate which modulates transmissibility throughout the year. As a result, many respiratory viruses show clear seasonal variation in prevalence; the most well-known example being seasonal influenza which peaks every winter in the temperate zone of the Northern Hemisphere (Petrova and Russell, 2018). A similar pattern is seen for the four seasonal human coronaviruses: HKU1, NL63, OC43 and 229E (hereafter collectively referred to as “seasonal CoVs”) (Al-Khannaq *et al*., 2016; Friedman *et al*., 2018; Galanti *et al*., 2019; Góes *et al*., 2019; Huang *et al*., 2017; Killerby *et al*., 2018). These viruses cause respiratory infections which usually are mild and primarily affect young children.

Previous influenza pandemics have swept the world in multiple waves often but not always coinciding with winter months in temperate climates (Amato-Gauci *et al*., 2011; Taubenberger *et al*., 2019; Viboud *et al*., 2005, 2016). The 1968-1970 global influenza pandemic was sparked by a new influenza A/H3N2 virus, with a divergent hemagglutinin protein, which replaced the A/H2N2 virus circulating for 10 years previously (Viboud *et al*., 2005). The virus spread rapidly, but the viral dynamics and mortality was not synchronized across countries. While many experienced two more-severe flu seasons in the winters of 1969 and 1970, the US had higher mortality in the first season, while European countries, Japan, and Australia, had higher mortality in the second, and Canada had roughly equal mortality in both (Viboud *et al*., 2005).

The 2009 pandemic H1N1 virus (A/H1N1pdm09) originated in March 2009 in Mexico and spread around the globe within weeks. Only a few European countries saw substantial circulation of H1N1pdm09 in the spring of 2009. Instead the virus showed low prevalence over the summer and pronounced peaks in the following autumn and winter in many countries (Amato-Gauci *et al*., 2011). A/H1N1pdm09 has subsequently transitioned into a seasonal pattern causing winter epidemics in temperate climates.

Here we use data on seasonal variation in prevalence of seasonal CoVs in Sweden and model the impact of this variation on the possible future spread of SARS-CoV-2 in the temperate zone of the Northern Hemisphere. We also explore different scenarios of SARS-CoV-2 spread in temperate and tropical regions and show how variation in epidemiological parameters affects a potential pandemic and the possibility of transitioning to an endemic state.

## l. SEASONAL CORONAVIRUS PREVALENCE

Data on seasonal variation of HKU1, NL63, OC43 and 229E diagnoses in respiratory samples was obtained from the routine molecular diagnostics at the Karolinska University Hospital, Stockholm, Sweden. The laboratory provides diagnostic services to six of seven major hospitals and approximately half of outpatient care in the Stockholm county (2.2 million inhabitants). We extracted pseudonymized data on all analyses for the four viruses between Jan 1, 2010 and Dec 31, 2019. The dataset included a total of 52,158 patient samples with 190,257 diagnostic tests, of which 2,084 were positive for any of the coronaviruses (229E = 319; NL63 = 499; OC43 = 604; HKU1 = 355; OC43/HKU1 = 307). Metadata included information about date of sampling and age of patient. In the period of Jan 1, 2010 to Nov 5, 2017, the coronavirus diagnostic was done using in-house assays (Tiveljung-Lindell *et al*., 2009). From Nov 6, 2017 to Dec 31, 2019, samples were analysed using the commercial kit Allplex Respiratory Panels (Seegene Inc., Seoul (South Korea)). This commercial kit does not distinguish between HKU1 and OC43, and for this reason positive tests for these two viruses were combined for the entire study period.

The fraction of tests that were positive for the four seasonal CoVs showed a strong and consistent seasonal variation, see Fig. 1. From December to April approximately 2% of tests were positive, while less than 0.2% of tests were positive between July to September, i.e. a 10-fold difference (Fig. 1, right). The strength of variation of the transmission rate through the year could be of high relevance to the spread of SARS-CoV-2 in 2020 and following years.

**FIG 1.**
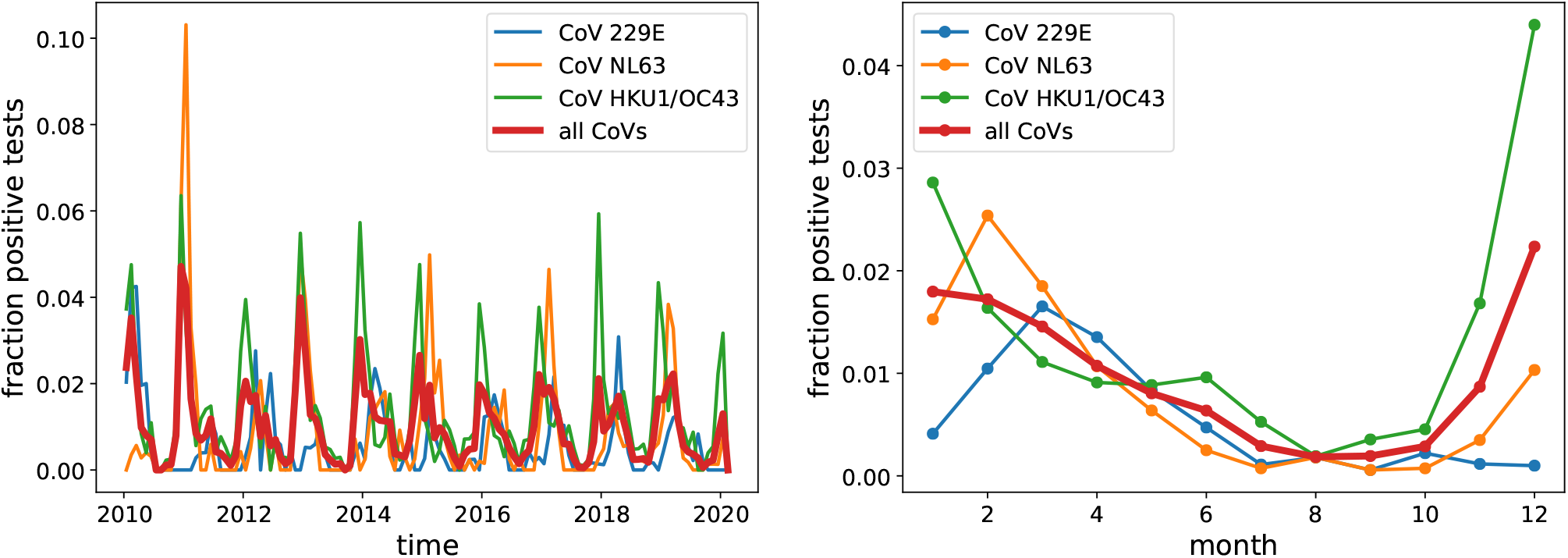
Seasonal variation in the fraction of positive CoV tests in Stockholm, Sweden. The left panel shows test results between 2010 and 2019. The right panel shows aggregated data for all years. All CoVs show a marked decline in summer and autumn, with HKU1/OC43 peaking January–December, and NL63 and 229E peaking in February–March.

## ll. BASIC MODEL

We consider simple SIR models (Kermack and McKendrick, 1991) with an additional category *E* of exposed individuals of the form

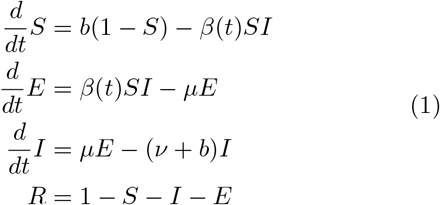

where *β*(*t*) is the rate at which an infected individual infects a susceptible one, *µ* is the inverse incubation time, *ν* is the recovery rate and *b* is the population turn-over rate. Depending on the analysis below, we implement several such populations that exchange individuals through migration, for details see Supplementary Methods. Stochasticity is implemented through Poisson resampling of the population once every serial interval *µ*^-1^ + (*ν* + *b*)^-1^. The population turnover *b* rate is immaterial for a pandemic scenario, but important for our analysis of seasonal CoVs, and should be interpreted as the sum of the birth rate and the rate at which previously immune individuals become susceptible due to immune waning and escape. We review general properties of such model in the Supplementary Materials. Following previous work, we parameterize transmissibility as

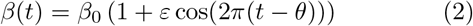

where *β*_0_ is the average annual infection rate, *ε* is the amplitude of seasonal forcing which modulates transmissibility through the year, and *θ* is the time of peak transmissibility (Chen and Epureanu, 2017; Dushoff *et al*., 2004). For simulations of the pandemic, we will add an additional term to *β*(*t*) that accounts for infection control measures in heavily affected areas, see Supplementary Materials.

## lll. MODEL PARAMETERIZATION USING SEASONAL COV OBSERVATIONS

Seasonal CoVs are endemic throughout the world and we therefore expect that viruses are imported throughout the year. We model this import through migration of susceptible individuals with rate *m* that return exposed with probability *x*. Humans develop immune responses to CoVs rapidly and subsequent challenge studies show reduced susceptibility and less severe disease for a year (Callow *et al*., 1990). Antibodies against SARS-CoV-1 persist for several years (Guo *et al*., 2020). This is consistent with the observation that about 50% of all positive samples in our data come from patients older than 10 years with a flat distribution across age groups. In analogy to the attack rate of seasonal influenza, we assume humans suffer from a seasonal CoV infection on average every 10 years (*b* = 0.1*/y*). Furthermore, we use ⟨*R*_0_⟩ = 2.3, a recovery rate of 0.2days^-1^, and an incubation period of 5 days.

With these assumptions, we can solve the model and compare the resulting trajectories to the seasonal variation in prevalence of seasonal CoVs, see Fig. 2. In Stockholm, seasonal variation of CoVs (especially HKU1/OC43) is very consistent across years (see Fig. S3). We therefore fit the SIER model to the average seasonal variation across years by calculating the squared deviation of observed and predicted prevalence relative to their respective mean values. Simulations of the model are compatible with observations in two separate regions of parameter space: If Northern Europe was very isolated with less than 1 in 1,000 susceptible individuals returning with a seasonal CoV infection from abroad each year, weak seasonality of around *ε* = 0.15 would be sufficient to generate strong variation through the year compatible with observations (Fig. 2, bottom-left ridge). In this regime, prevalence is oscillating intrinsically with a period that is commensurate with annual seasonal oscillations giving rise to a resonance phenomenon with annual or biennial patterns even for weak seasonal forcing (Chen and Epureanu, 2017; Dushoff *et al*., 2004).

**FIG 2.**
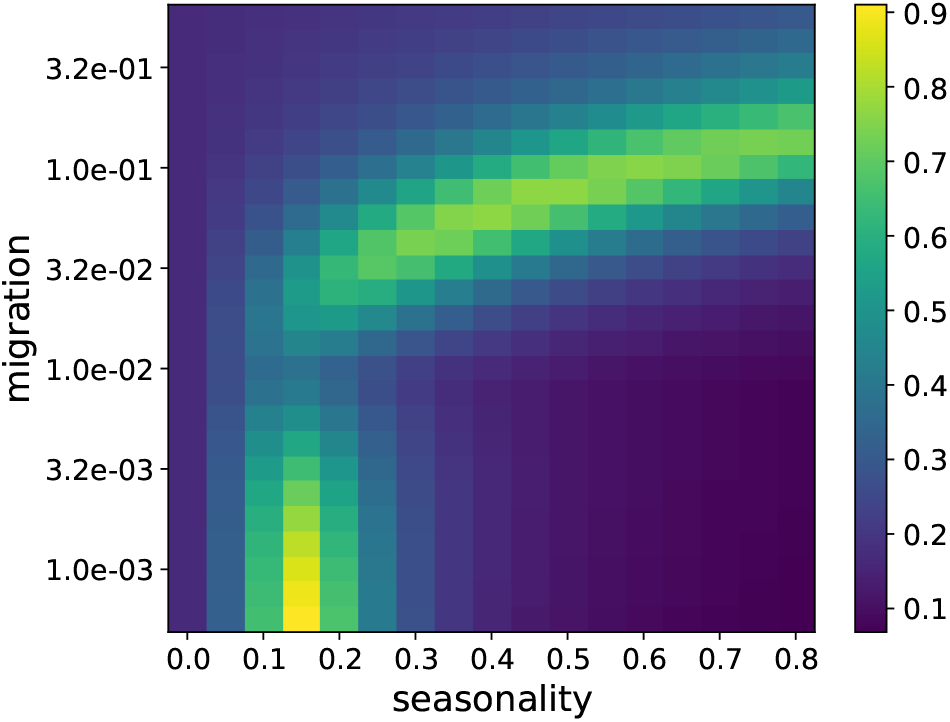
Compatibility of SIER model trajectories with observations. The heatmap shows the inverse mean squared deviation between the model trajectories and the observed seasonal forcing in seasonal CoV prevalence. Model and observations are compatible (yellow shading) in a region of parameter values corresponding to low migration/weak seasonality and second region at high migration/strong seasonality. Migration refers to the rate per year of a susceptible to return from a abroad with a CoV infection.

If the rate of import of seasonal CoV infections is higher, imports dampen the resonance and much stronger seasonality, with values between *ε* = 0.3 and 0.7, is required to fit the observations (Fig. 2, top-right-and-center ridge). In this regime, seasonal variation in transmissibility modulates the size of micro-outbreaks triggered by imported cases in a mostly immune population.

These two scenarios differ slightly in the time of year at which peak transmissibility *θ* occurs: When transmission is mostly local and seasonality is amplified by resonance, *θ* needs to be around October–November to fit the data with most cases in December–January. In the second scenario with high connectivity, *θ* needs to be in December–January coinciding with the peak in prevalence. Given that most countries are highly connected, we focus here on exploring the high-import and strong seasonal forcing scenario. This scenario, with maximal *β* in mid-winter, is also more compatible with climate variation around the year. The qualitative behavior of the fit is robust to uncertainty in *R*_0_ and the frequency of reinfection *b*.

## lV. SCENARIOS FOR SARS-COV-2 PANDEMICS IN 2020 AND 2021

The analysis of seasonal CoV prevalence patterns allowed us to constrain parameter ranges and explore different scenarios of SARS-CoV-2 spread around the globe, in particular in temperate climates like Northern Europe. Here we explore scenarios where temperate regions have a seasonal forcing of between *ε* = 0.3 and 0.7 and migration rates of 0.01/year. Early estimates suggest an incubation time of about 5 days and an average serial interval of 7-8 days (Wu *et al*., 2020). Our model uses an average incubation time of 5 days (Backer *et al*., 2020) and an infectious period of 5 days.

To match the *R*_0_ estimates for the early outbreak with our parameterization of transmissibility in Eq. 2 we need to account for the fact that December/January are winter months in Hubei and peak transmissibility in Hubei likely corresponds to *θ ≈* 0 (0 being the beginning of the year, so a *θ* in December/January). An *R*_0_ ≈ 3 in winter in Hubei and a seasonal forcing of *ε* = 0.4 implies an annual average ⟨*R*_0_⟩ = *β*_0_*/ν* = 2.2. This reasoning leads to our parameter choice of *β*_0_ = 158/year, *ν* = 72/year, *θ* = 0. We assume the outbreak started at *t* = 2019.8 in Hubei with one infected individual and use *N* = 6 × 10^7^ as population size. To incorporate infection control measures, transmissibility is reduced by 50% once prevalence reached 3% (third order Hill-function, see Supplemental Material). Introductions to a location like Northern Europe with *ε* = 0.5 (*i*.*e*. slightly stronger seasonal forcing then Hubei) are assumed to happen at a rate of 0.01 per year for each infected individual elsewhere. The simulation of the SIER model in different regions is deterministic, but migration is implemented stochastically by Poisson resampling of the average number of migrating individuals. Fig. 3 shows simulated trajectories of SARS-CoV-2 prevalence in the temperate Northern Hemisphere assuming the outbreak started in Hubei early December 2019. Depending whether the peak transmissibility of SARS-CoV-2 in the northern temperate zone is in November, January, or March, the simulation predicts a main peak in the first half of 2020, a main peak in winter 2020/2021, or two similarly sized peaks.

**FIG 3.**
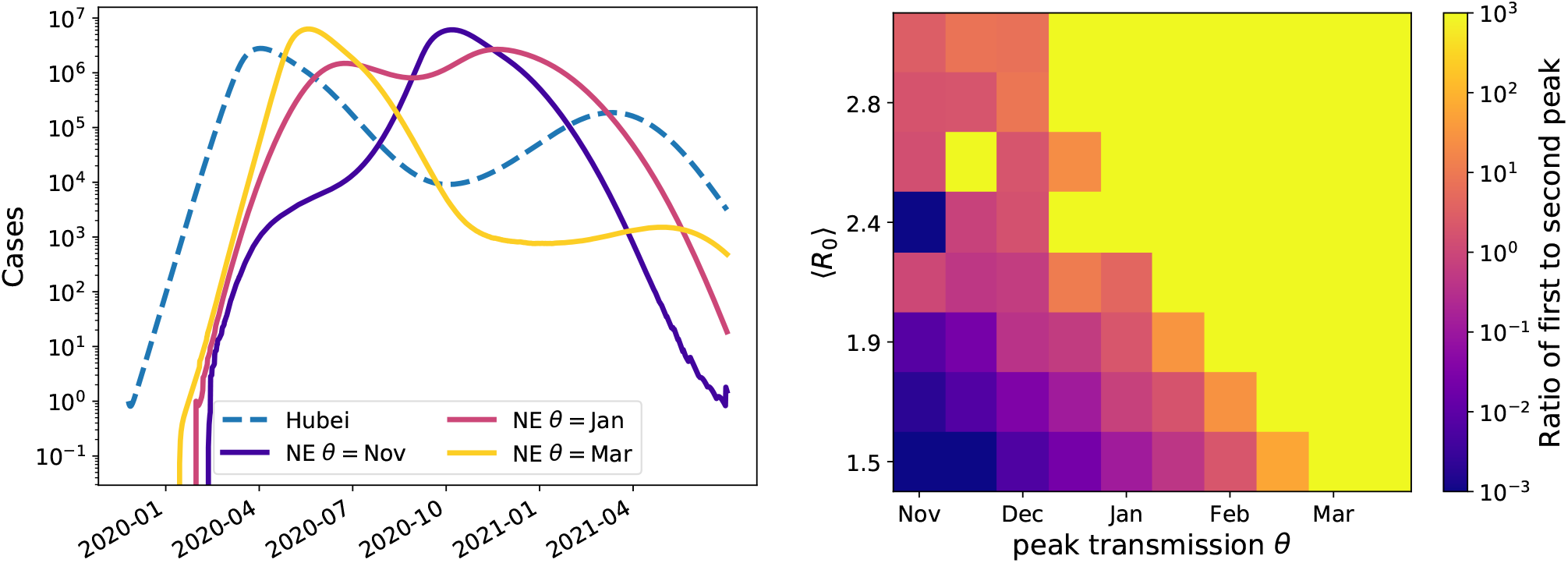
Model predictions for SARS-CoV-2 case numbers in temperate zones for a pandemic scenario. The left panel shows example trajectories assuming SARS-CoV-2 transmissibility peaks in November, January, or March. These outbreaks in Northern Europe (‘NE’) are assumed to be seeded by the outbreak in Hubei (model trajectory shown as a dashed line). Within the model, these cases are exported at rate of 0.01/year to temperate Northern Europe with an average *R*_0_ = 2.2 and seasonal forcing of *ε* = 0.5. Corresponding graphs for different values of *R*_0_ and the migration rate are shown in Supplemental Fig. S4. The right panel shows the ratio of the first and second peak for a range of different combinations of *R*_0_ and *θ*. The yellow area corresponds to parameter combinations with essentially only an early peak similar to the yellow line on the left. The blue/purple area shows parameter combinations for which a peak in late 2020 dominates, as with the purple line on the left, while the central pink/orange band shows the combinations giving rise to two comparable peaks. These simulations are for *ε* = 0.5. Similar results were obtained for *ε* = 0.3 and 0.7, see Supplementary Fig. 6.

To explore possible scenarios more systematically, we ran such simulations for a range of values for *R*_0_ and peak transmissibility *θ* and recorded whether we observe and early peak, a late peak, or a two peaks. The right panel of Fig. 3 shows the ratio of the height of these peaks for different values. Rapid growth (high *R*_0_) and late transmission peaks result in a large peak in the first half 2020, while lower *R*_0_ and transmission peaks in early winter favor a large secondary peak. These two scenarios are separated by a band of parameter values that give rise to two pandemic waves in the winters of 2020 and 2021 in the Northern Hemisphere. Individual trajectories for a variety of parameter combinations are given in Fig. S4. The qualitative behavior is robust to model perturbations and parameter variation as long as seasonal forcing is strong. With weak forcing (*ε* = 0.15), the model predicts a single peak for most combinations of *R*_0_ and migration rates (see Fig. S5).

The uncertainty in parameter values and the potential impact of infection control measures imply that all scenarios are plausible and should be considered when developing pandemic prevention and containment strategies.

## V. GLOBAL PROJECTIONS

In absence of control measures, outbreaks initially grow exponentially within well-mixed communities, and at a certain rate the virus will be carried to other regions and potentially seed new outbreaks. Such export to new locations is initially unlikely, but becomes next to certain once the outbreak size exceeds the inverse probability that a given individual migrates while infected. We have witnessed such rapid dispersal to SARS-CoV-2 to many countries across the globe during January and February 2020.

Every location has a different socio-economic profile such that the growth rate of the epidemic (and hence *R*_0_) might differ. The superposition of many such subpopulations with a range of *R*_0_ values and seasonal variation in transmission will result in dynamics that are qualitatively different from a single population SIR model. In particular, such variation result in a pandemic spread out over 2 years before the virus possibly becomes endemic.

Fig. 4 shows the result of such a simulation of 1,000 populations. Populations were divided between northern temperate (50%), southern temperate (10%), and tropical (40%) and assigned parameters as follows:

**FIG 4.**
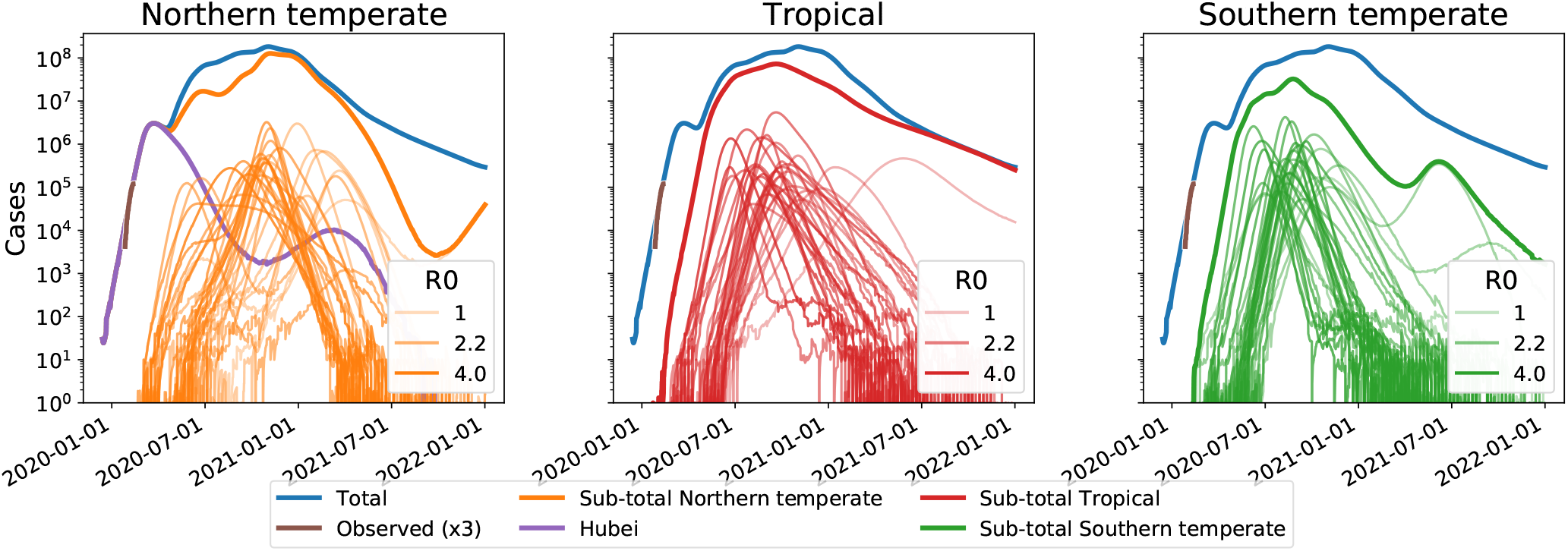
Extended circulation through overlapping epidemics in variable subpopulations. These simulations of a pandemic scenario assume 1,000 sub-populations with an average *R*_0_ of 2.2 and standard deviation 0.5, 40% of which have weak seasonal forcing *ε* [0, 0.2] (tropical) and the remainder have strong variation with *ε* [0.25, 0.75]. The super-position of many variable epidemics can result in a global prevalence that decays only slowly through 2020 and 2021. Lighter lines have lower *R*_0_, darker lines have higher *R*_0_. The actual observed case counts reported for Hubei are added (brown line) and multiplied by three to account for possible under-reporting of mild cases. A subset of 30 randomly chosen simulations are plotted for each region. Analogous figures for different *R*_0_ parameter values are shown in Supplementary Fig. 7.

- ⟨*R*_0_⟩ was drawn from a normal distribution with mean 2.2 (see Supplementary Fig. 7 for ⟨*R*_0_⟩ = 1.5 and 3.0) and standard deviation 0.5.
- Seasonal forcing *ε* was drawn from a uniform distribution between 0.25 and 0.75 for temperate regions, between 0 and 0.2 for tropical regions.
- Peak transmissibility *θ* of temperate regions was drawn from normal distributions with standard deviation 0.1 and peak at 0 for northern regions and 0.5 for southern regions. *θ* for tropical regions was chosen uniformly from between 0 and 1.
- Population sizes were drawn from a log-normal distribution with *σ* = 1 and a mean such that all populations sum to 7.6 billion.
- Migration rates were sampled from a log-normal distribution with *σ* = 1 and a mean of 0.01.

For Hubei, we use the same parameters as described in section IV.

The variation in *R*_0_ and migration rate result in a super-position of fast and slow epidemics seeded at different times. The initial phase is dominated by fast epidemics driving rapid dispersal, in particular in the tropics, while slow epidemics dominate later in 2020 and 2021. With the parameter setting used in Fig. 4, the Northern temperate regions see most circulation in winter 2020/2021. In accordance with Fig. 3, this peak shifts more towards early 2020 for higher *R*_0_, see Supplementary Fig. 7.

After several years, SARS-CoV-2 could become a seasonal CoV with characteristic winter outbreaks as shown in Fig. 1. Such a scenario is demonstrated in Fig. 5 where a simulation similar to the one shown in Fig. 4 is run for 12 years, with the added assumption that after infection an individual become susceptible to SARS-CoV-2 again at a rate of 0.1 per year as we assumed for seasonal CoV above. After a pronounced low in 2020-2024, prevalence recovers and settles into a seasonal pattern, similar to that of the four existing seasonal CoVs.

**FIG 5.**
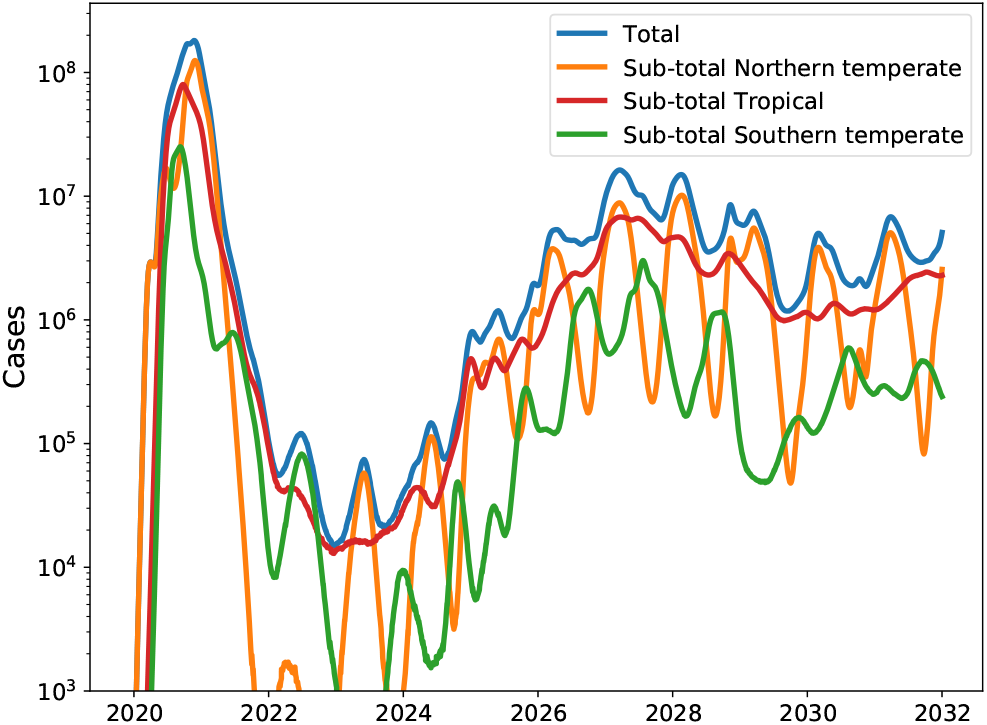
Transition to an endemic seasonal virus. If previously infected individuals can be reinfected after some time, as for example by seasonal influenza virus, SARS-CoV-2 could develop into a seasonal CoV that returns every winter. This would typically happen at much lower prevalence than peak pandemic levels. These simulations assume reinfection on average every 10 years.

## Vl. DISCUSSION

We report on the possible influence of seasonal variation on the spread of SARS-CoV-2 in the Northern Hemisphere, in a pandemic scenario. We find that seasonal variation in transmissibility has the potential to modulate the spread of SARS-CoV-2 with a wide range of possible outcomes that need to be taken into account when interpreting case counts and projecting the outbreak dynamics. The onset of spring and summer could, for example, give the impression that SARS-CoV-2 has been successfully contained, only for infections to increase again in 2020-2021 winter season. Even in Hubei virus circulation might decrease due to containment measures and the arrival of spring but might increase again towards the end of the year. Whether a pandemic in the temperate regions of the Northern Hemisphere would peak early in 2020, late in 2020, or show multiple waves as H1N1pdm did in 2009, depends on the timing of peak transmissibility and the rate of spread (*R*_0_ and serial interval).

This study is meant as an exploration of how such a pandemic could unfold, not as a prediction of any particular scenario. The results we present are critically dependent on the assumptions i) that the outbreak will develop into a pandemic, ii) that the transmissibility of SARS-CoV-2 shows seasonal variability of sufficient strength (range *ε* = 0.3 to 0.7), and iii) that parameters like *R*_0_ estimated from the early phase of the outbreak are comparable in other populations.

These assumptions are not implausible but not certain: cases of SARS-CoV-2 has been in several countries in Asia apart from China, as well as in Europe, Africa, North America, and Australia (WHO Emergency Committee, 2020b), and mild or asymptomatic cases make detection and thus prevention of spread by isolation challenging, *e*.*g*. airport screening as a preventive measure is unlikely to prevent spread and local seeding (Quilty *et al*., 2020). Person-to-person transmission of the virus has been documented in several countries outside of China, including large outbreaks in Iran, South Korea, and Italy (European Centre for Disease Prevention and Control (ECDC), 2020). It is likely that not all exported cases have been detected, and some may have seeded outbreaks outside of China that have yet to be detected.

The seasonal CoVs show a strong and consistent seasonal variation, and modeling suggests that this requires strong variation in transmissibility throughout the year. It should be noted, however, that SARS-CoV-2 does seem to transmit in tropical climates like Singapore, and so winter is not a necessary condition of SARS-CoV-2 spread. Furthermore, our models are compatible with work by Luo *et al*. (2020) showing that recent trends in different regions across East-Asia imply that seasonality alone is unlikely to end SARS-CoV-2 spread. Precise values for the underlying model parameters and the effect of infection control measures are currently unavailable. For this reason we explored a range of parameter values to assess the robustness of the results to model assumptions.

The simulations presented here are scenarios that emerge from simplified abstract models, but they nevertheless demonstrate that a wide variety of outcomes are compatible within the limits of the current knowledge about the outbreak. The implications of our work are that: 1) reductions in prevalence need not be attributable to successful interventions, but could be due to seasonal variation in transmissibility, 2) sub-population dynamics can differ greatly, meaning that case count trajectories in one country should be used cautiously to inform projections in a second country, even in the same climate zone, 3) seasonal variation might slow down a pandemic and thereby provide a window of opportunity for better preparation of health care systems world-wide by scaling up capacity for care and diagnostics, and potentially through rapid development of antivirals and vaccine, and 4) after several years SARS-CoV-2 could develop into an endemic seasonal CoV similar to the transition of the 2009 A/H1N1 pandemic influenza virus into a seasonal influenza virus.

The overall impact of a potential SARS-CoV-2 pandemic depends critically on the case fatality ratio (CFR), which we have not modelled here. At present, uncertainty in the CFR is high due to likely over-representation of severe cases in the statistics and a delay between diagnosis and recovery/death (Battegay *et al*., 2020). Even with this unknown, seasonal variation in transmissibility of SARS-CoV-2 and underlying differences in migration, introduction times, and attack rate should thus be taken into account when monitoring and projecting global transmission, planning further surveillance of the epidemic, and developing pandemic prevention and containment strategies.

## Data Availability

All relevant data and script that generate the graphs are available in a dedicated github repository at github.com/neherlab/CoV_seasonality.

## Vll. CODE AND DATA AVAILABILITY

All relevant data and script that generate the graphs are available in a dedicated github repository at github. com/neherlab/CoV_seasonality.

### ACKNOWLEDGEMENTS

We are grateful to all physicians, scientists, and public health authorities globally for sharing case numbers and early research findings without delay. We would like to thank Christian Althaus for critically reading the manuscript and providing timely and valuable feed back. This research was supported in part by the National Science Foundation under Grant No. NSF PHY-1748958 (RAN during his visit to KITP).

## AUTHORS’ CONTRIBUTIONS

JA conceived the idea of the study. RAN developed the model and simulations. JA and RD provided the data on seasonal CoVs. RAN, EBH and VD performed the modeling and simulation, and generated the graphs. All authors interpreted the data, drafted and commented on the manuscript, and contributed to the final version.

